# Quantification of Cerebrospinal Fluid Tumor DNA in Lung Cancer Patients with Suspected Leptomeningeal Carcinomatosis

**DOI:** 10.1101/2024.01.03.23300646

**Authors:** Tej D. Azad, Shigeki Nanjo, Michael C. Jin, Jacob J. Chabon, David M. Kurtz, Aadel A. Chaudhuri, Ian D. Connolly, Angela Bik-Yu Hui, Chih Long Liu, David Merriott, Ryan Ko, Christopher Yoo, Justin Carter, Emily Chen, Rene Bonilla, Akito Hata, Nobuyuki Katakami, Kei Irie, Seiji Yano, Ross Okimoto, Trever G. Bivona, Aaron M. Newman, Michael Iv, Seema Nagpal, Melanie Hayden Gephart, Ash A. Alizadeh, Maximilian Diehn

## Abstract

**Introduction:** Cerebrospinal fluid tumor-derived DNA (CSF-tDNA) analysis is a promising approach for monitoring neoplastic processes of the central nervous system. We hypothesize that analysis of CSF-tDNA in patients with advanced lung cancer improves the sensitivity of leptomeningeal disease (LMD) diagnosis and enables central nervous system response monitoring.

**Methods:** We applied CAPP-Seq using a lung cancer-specific sequencing panel to 81 CSF, blood, and tissue samples from 24 patients with advanced lung cancer who underwent lumbar puncture (LP) for suspected LMD. A subset of the cohort (N = 12) participated in a prospective clinical trial of osimertinib for refractory LMD in which serial LPs were performed before and during treatment with.

**Results:** CSF-tDNA variant allele fractions (VAFs) were significantly higher than plasma circulating tumor DNA (ctDNA) VAFs (median CSF-tDNA, 32.7%; median plasma ctDNA, 1.8%; *P* < 0.0001). Concentrations of tumor DNA in CSF and plasma were positively correlated (Spearman’s ρ, 0.45; *P* = 0.03). For LMD diagnosis, cytology was 81.8% sensitive and CSF-tDNA was 91.7% sensitive. CSF-tDNA was also strongly prognostic for overall survival (HR = 7.1; *P* = 0.02). Among patients with progression on targeted therapy, resistance mutations, such as *EGFR* T790M and *MET* amplification, were common in peripheral blood but were rare in time-matched CSF, indicating differences in resistance mechanisms based on anatomic compartment. In the osimertinib cohort, patients with CNS progression had increased CSF-tDNA VAFs at follow up LP. Post-osimertinib CSF-tDNA VAF was strongly prognostic for CNS progression (HR = 6.2, *P* = 0.009).

**Conclusions:** Detection of CSF-tDNA in lung cancer patients with suspected LMD is feasible and may have clinical utility. CSF-tDNA may improve the sensitivity of LMD diagnosis, enable improved prognostication, and drive therapeutic strategies that account for spatial heterogeneity in resistance mechanisms.

## Introduction

Metastasis of malignant cells to the leptomeninges, cerebrospinal fluid (CSF) compartment, and subarachnoid space results in leptomeningeal disease (LMD).^1^The prognosis of LMD is poor, with only 10% of solid tumor patients surviving beyond one year.^2–5^ The advent of oncogene-directed therapies for patients with non-small-cell lung cancer (NSCLC) LMD has extended median survival from one month up to nearly one year.^1, 6–8^ Currently, diagnosis and response assessment in LMD relies upon CSF cytology, the clinical standard, as well as physical examination and magnetic resonance imaging (MRI) of the brain and spine.^9^ CSF cytology is highly specific for LMD, but has a reported sensitivity of only 50-60%.^1, 9–11^

Detection of tumor-derived DNA in CSF (CSF-tDNA) has emerged as a novel method for detecting and monitoring neoplastic processes of the central nervous system,^12–18^ applying principles of peripheral blood-based circulating tumor DNA (ctDNA) detection to CSF. Many studies investigating the role of CSF-tDNA in LMD have been case reports,^16, 19–22^ and comprehensive studies with prospective cohorts remain rare.^23–28^ The increasing number of potential genetic drivers in advanced-stage NSCLC prompted the development of next-generation sequencing (NGS)-based multiplex plasma ctDNA assays for the non-invasive detection of a wide range of genomic events. Recent cohort studies have demonstrated the feasibility of detecting CSF-tDNA in patients with *EGFR*-mutant and *ALK*-rearranged NSCLC.^27–37^ Consistent observations across these reports are higher variant allele fractions (VAFs) in CSF compared to plasma and better concordance between the original tumor and CSF-tDNA, than between tumor and plasma ctDNA. Preliminary results suggest the sensitivity of CSF-tDNA is superior to that to CSF cytology^23^ and the utility of CSF-tDNA to predict treatment response remains unknown.

In the current study, we aimed to develop a CSF-tDNA assay using CAPP-Seq,^38–42^ a targeted NGS-based method originally developed for analysis of plasma ctDNA, for the diagnosis and monitoring of LMD in patients with lung cancer. We hypothesized that detection of CSF-tDNA in patients with advanced lung cancer improvs the sensitivity of LMD detection, enables analysis of treatment resistance mutations, and allows monitoring of LMD response to targeted therapy (**Fig. 1**).

**Figure 1.**
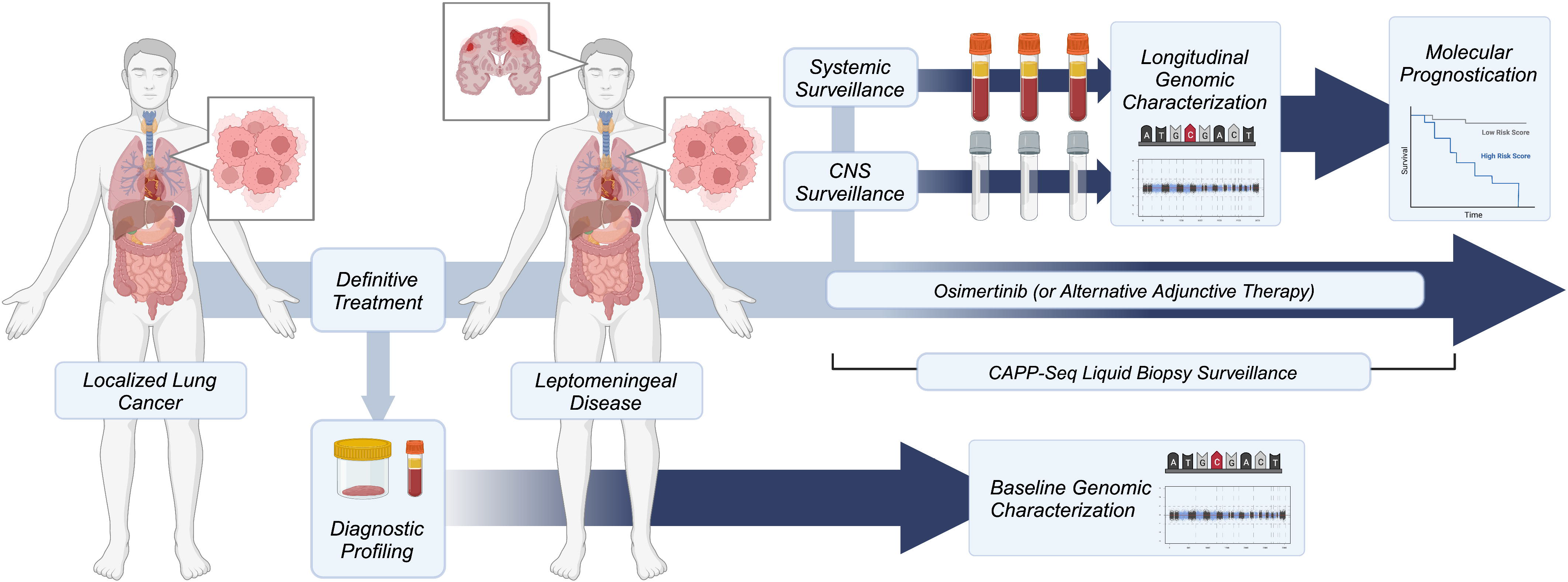
Schematic overview of study and proposed future clinical integration of CAPP-Seq informed LMD management.

## Results

### Detection of CSF-tDNA in lung cancer patients with suspected LMD

We profiled 81 cerebrospinal fluid (CSF), blood, and tissue samples from 24 total patients with advanced lung adenocarcinomas who underwent lumbar puncture for evaluation of leptomeningeal disease (LMD, **Fig. S1**). Median age of the patients in our cohort was 64 (IQR, 55.3-72.0). The majority of our cohort was female (N = 18) and did not have a history of smoking (N = 16). Most patients had concurrent brain metastases (N = 17) at time of LMD evaluation. Our cohort was comprised of 20 patients with EGFR-mutant tumors, two patients with ALK-driven tumors, and two patients with non-EGFR and non-ALK mutated lung cancer (**Fig. 2A**). Eighteen patients had definitive LMD and six patients had possible LMD (see **Methods** for definitions). CSF-tDNA variant allele fractions (VAFs) were significantly higher than plasma ctDNA VAFs (median CSF-tDNA, 32.7%; median plasma ctDNA, 1.8%; *P* < 0.0001; **Fig. 2B**). We further observed a significant positive correlation between CSF-tDNA VAF and plasma ctDNA VAF (Spearman’s ρ, 0.45; *P* = 0.03; **Fig. 2C**).

**Figure 2.**
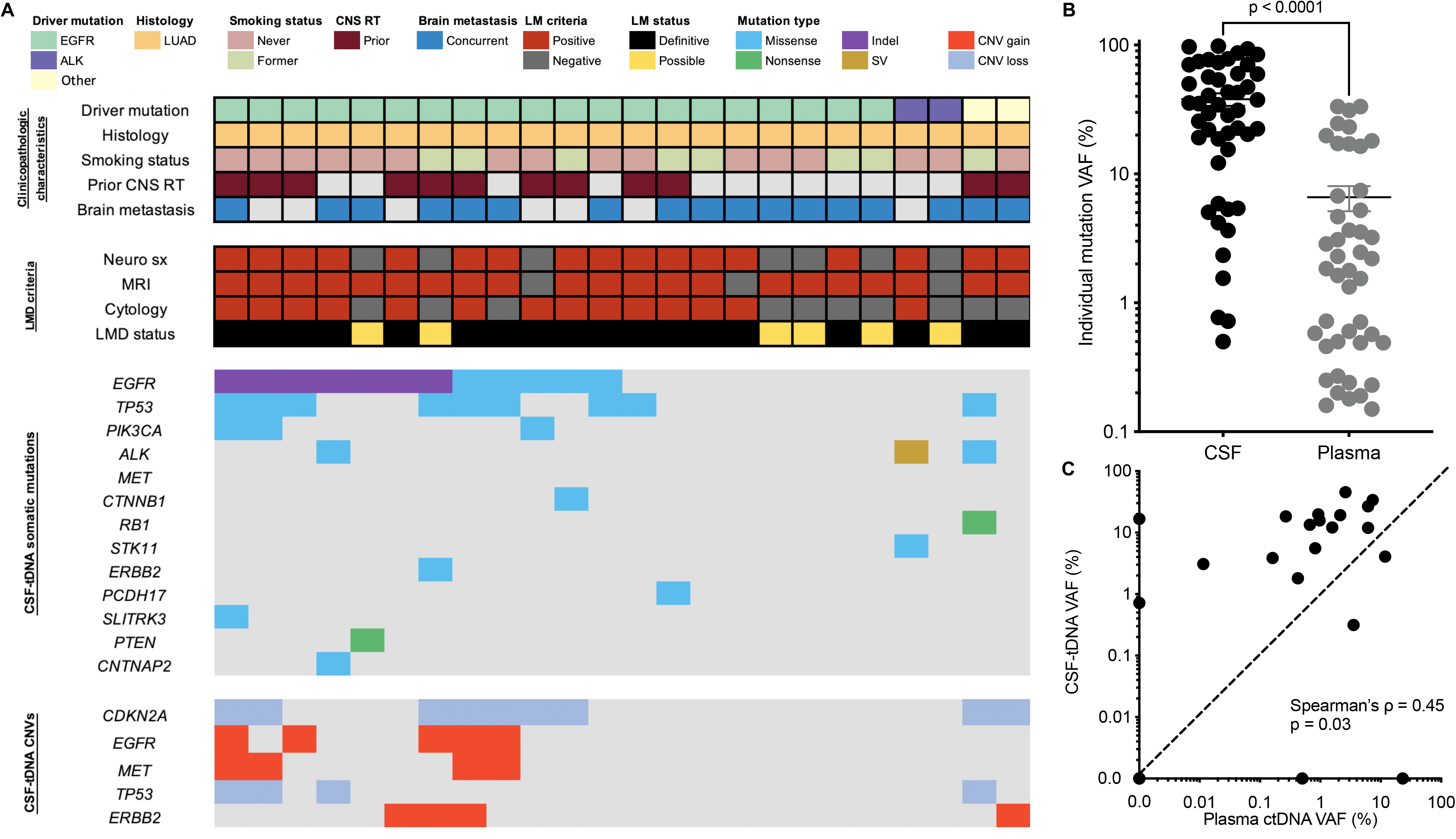
Detection of CSF-tDNA in lung cancer patients with suspected LMD. (**A**) Clinical characteristics and CSF-tDNA detection results. Each column represents a patient and each row a parameter (*e.g*. driver mutation). Similarly, the associated co-mutation plot depicts patient-level mutational profiles of CSF-tDNA in patients with lung cancer genotyped by our lung cancer-specific NGS panel. (**B**) Comparison of individual mutation VAFs in CSF-tDNA and in plasma ctDNA. P value calculated by the Mann-Whitney test. (**C**) Correlation of CSF-tDNA VAF with plasma ctDNA VAF. P value and ρ were calculated by Spearman correlation.

### Comparison of CSF-tDNA to cytology and imaging for diagnosis of LMD

Classically, detection of malignant cells in CSF by cytopathology is considered the gold standard for LMD diagnosis.^43^ Of 12 patients with positive CSF by cytopathology or by a clinical *EGFR* PCR assay, CSF-tDNA was positive in 11 (92%). To account for this, we compared sensitivity of cytology, MRI, and CSF-tDNA in detecting definitive LMD (see **Methods).** In our cohort, we found that cytology was 81.8% sensitive for diagnosis of definitive LMD while MRI was only 80.0% sensitive. In contrast, CSF-tDNA was 91.7% sensitive for definitive LMD (**Fig. 3A**).

**Figure 3.**
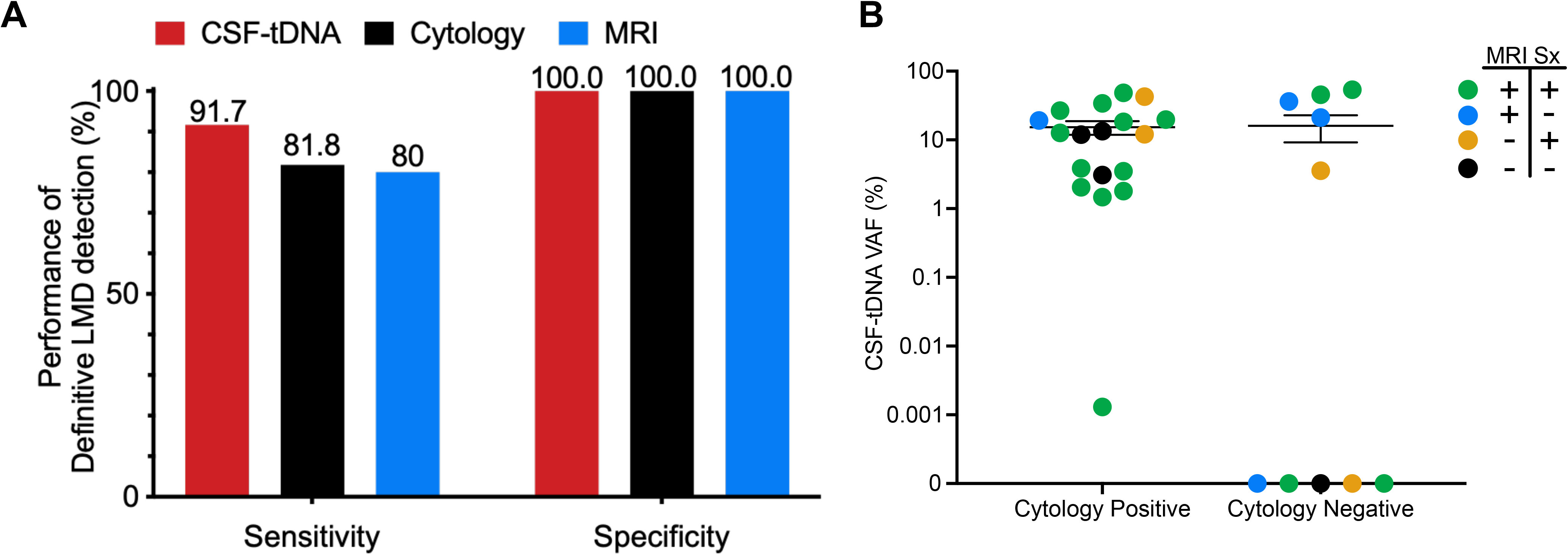
Performance of CSF-tDNA in diagnosis of definitive LMD. (**A**) Sensitivity of CSF-tDNA, cytology, and MRI in diagnosis of LMD. (**B**) Comparison of CSF-tDNA VAF between cytology-positive and cytology-negative patients, stratified by which criteria were met in the definition of LMD. The cases who had negative cytology were diagnosed by MRI of brain or spine with unequivocal evidence and progressive neurological symptoms consistent with LMD.

We next investigated the association between LMD and CSF-tDNA VAFs, including all samples with available CSF cytology, MRI, and history of neurological symptoms (N = 27). Mean CSF-tDNA VAF of cytology (+) samples (N = 18) was 15.3% while mean VAF of cytology (-) samples was 17.4% (N = 9, *P* = 0.43) (**Fig. 3B**). Importantly, 4/9 samples which were cytology (-) had detectable CSF-tDNA (mean VAF, 39.1%). Of these four samples, two had both MRI findings of LMD and progressive neurologic symptoms at the time of LP and two had MRI findings alone.

### Prognostic utility of CSF-tDNA for patients with suspected LMD

Next, we explored the prognostic value of CSF-tDNA detection in patients with LMD. We found that the presence of CSF-tDNA is strongly associated with poor OS (HR = 7.1; *P* =0.02; **Fig. 4**). In clinical CSF cytology, centrifugation of the sample is commonly performed to collect cells for further processing and the supernatant is generally discarded.^43^ CSF-tDNA isolation, as performed in this study, is compatible with cytology, as we centrifuged the CSF sample and extracted DNA from the supernatant. Thus, we tested the prognostic utility of considering both CSF cytology and CSF-tDNA. We found that patients who were CSF-tDNA and cytology positive had significantly worse overall survival (OS) than patients who were CSF-tDNA and cytology negative (HR = 8.4; *P* = 0.015; **Fig. S2A**). Four patients had discordant CSF-tDNA and cytology (CSF-tDNA positive, cytology negative). Prognosis for these patients appears to be better than patients with both CSF-tDNA and cytology positive but worse than patients who are CSF-tDNA and cytology negative (**Fig. S2B**).

**Figure 4.**
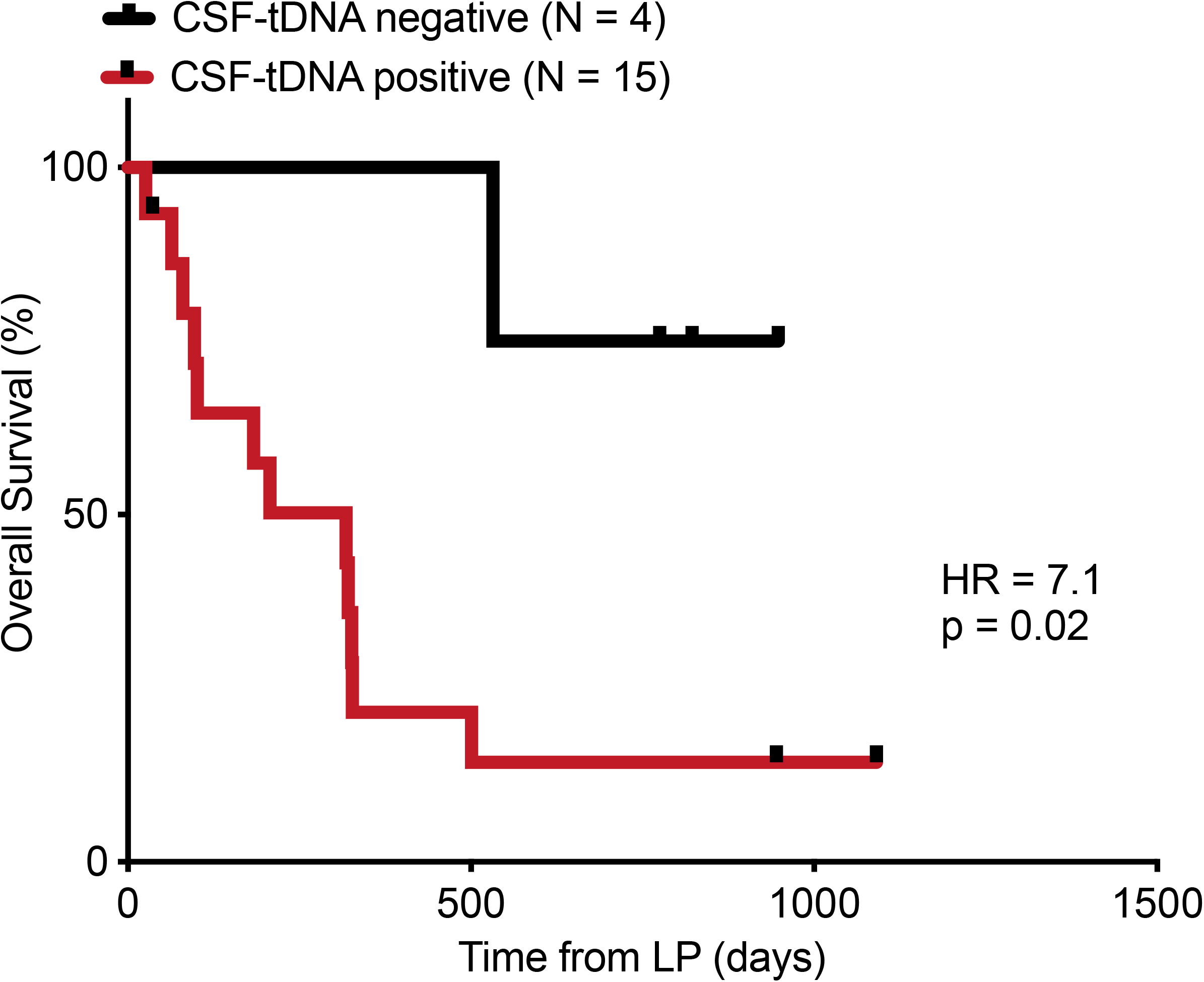
Association of CSF-tDNA with overall survival. Kaplan-Meier curve comparing patients with detectable (N = 15) and undetectable (N = 4) CSF-tDNA at first lumbar puncture for the endpoint of overall survival (*P* = 0.02, HR = 7.1 (95%CI, 2.3-21.9)). P value and hazard ratio were calculated from the log-rank test.

### Spatial heterogeneity of resistance mechanisms to targeted therapy

We next investigated the ability of time-matched CSF-tDNA and plasma ctDNA to detect resistance mechanisms to targeted therapy. In this analysis, we included EGFR-mutant lung adenocarcinoma patients who received EGFR tyrosine kinase inhibitor (TKI) therapy, had a pre-TKI sample available (CSF, N = 7; tumor biopsy, N = 1, pleural effusion, N = 1), and had available post-TKI therapy CSF and plasma samples, collected within two weeks of each other with no intervening treatment. To identify putative resistance mutations, we performed tumor-naïve variant calling^39^ on the post-TKI plasma sample, retaining mutations in genes known to be associated with resistance mutations (exonic SNVs, *EGFR, PIK3CA, KRAS, CDKN2A, RB1, ALK, KIT, MET*; copy number variants, *MET, ERBB2, EGFR*).^40^ Resistance mutations were defined as variants in these genes that were absent in the pre-TKI sample.

Seven patients (77%) in this analysis received osimertinib and two patients (22%) received erlotinib. Seven patients (77%) had CNS progression, one patient (11%) had non-CNS progression, and one patient (11%) did not have any progression after EGFR-TKI therapy. We detected putative resistance mutations in the peripheral blood of seven patients (77%), all of whom had CNS progression on EGFR-TKI therapy. Strikingly, resistance mutations were much less commonly observed in matched CSF though tumor DNA concentration was higher in CSF than plasma in the case (LUP112) where a shared resistance SNV was identified in both compartments. When we did detect emergent resistance mutations in the CSF (N = 2, *MET* amplification and *PIK3CA* SNV) the same mutations were also observed in matched plasma (**Fig. 5A**). No resistance mutations, in either plasma or CSF, were detected in the two patients without CNS PD.

**Figure 5.**
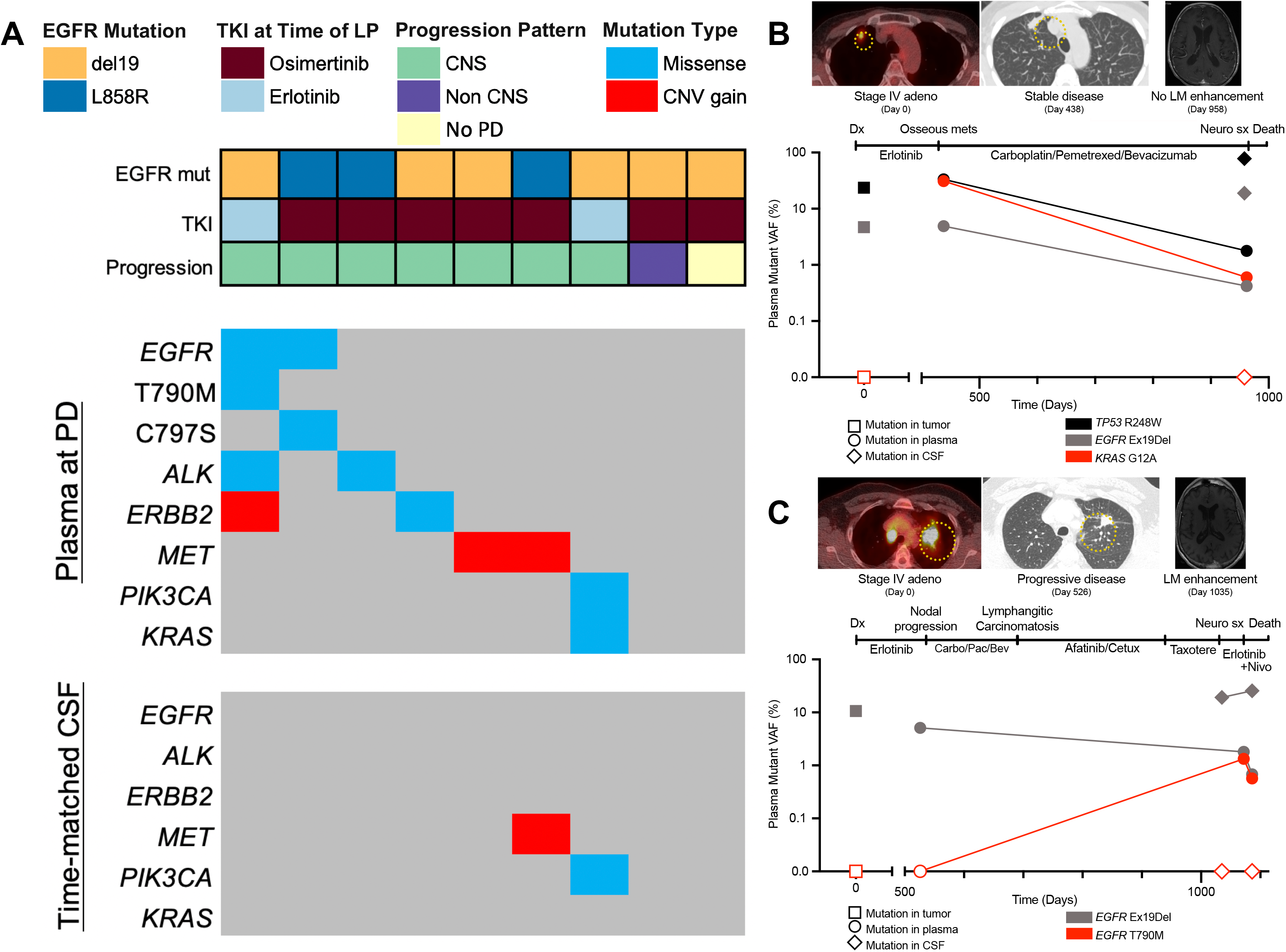
Emergent mechanisms of EGFR TKI resistance in plasma ctDNA and CSF-tDNA. This analysis was limited to EGFR-mutant lung adenocarcinoma patients who received EGFR-TKI therapy, had a pre-TKI sample available, and had available post-TKI therapy CSF and plasma samples, collected within two weeks of each other with no intervening treatment (N = 9). (**A**) Clinical characteristics co-mutation plot of resistance mutations. PD, progressive disease. (**B**) Patient (LUP112) with Stage IV adenocarcinoma with an emergent *KRAS* G12A mutation in plasma that was absent in the pre-treatment tumor. This mutation was absent in detected CSF-tDNA at time of LMD diagnosis. (**C**) Patient (LUP132) with Stage IV adenocarcinoma with an emergent *EGFR* T790M mutation and *ERBB2* amplification in plasma, both of which were absent in time-matched CSF.

We provide two illustrative cases to highlight these findings. In **Fig. 5B**, we present the case of a patient (LUP112) with Stage IV adenocarcinoma with known *EGFR* del19 and *TP53* R248W mutations. This patient was treated with erlotinib and developed osseous metastases. A plasma sample at this time revealed both mutations, but also a *KRAS* G12A mutation that had been absent in the initial tumor. The patient was transitioned to a regimen of carboplatin, pemetrexed, and bevacizumab but went on to later develop worsening headaches and altered mental status. Though MRI did not reveal leptomeningeal enhancement, a lumbar puncture was performed. Cytology was positive and CSF-tDNA was notable for high VAF detection of the original *EGFR* del19 and *TP53* R248W mutations. A plasma sample collected the same revealed these two mutations plus *KRAS* G12A, suggesting the resistant clone did not enter the CNS. In **Fig. 5C**, we present the case of another patient (LUP132) with Stage IV adenocarcinoma with a known *EGFR* del19 mutation, detected in both tumor and plasma. This patient went on to develop progressive headaches and had an MRI concerning for leptomeningeal enhancement. CSF-tDNA was detected in two consecutive lumbar punctures and *EGFR* del19 was detected in both. However, time-matched plasma samples revealed an emergent *EGFR* T790M mutation and emergent *ERBB2* amplification, both of which were absent in CSF.

### CSF-tDNA predicts response to osimertinib in a prospective cohort

A subset of our overall cohort was collected as part of a prospective study examining the efficacy of osimertinib for refractory LMD.^44^ Patients (N = 12) underwent lumbar punctures, before and 3 weeks after initiating treatment with osimertinib. Follow up lumbar punctures were accompanied by brain MRI and venipuncture for matched plasma ctDNA analysis. We found that patients with CNS progression had increased CSF-tDNA VAFs after three weeks of osimertinib (**Fig. 6A**). We next tested if CSF-tDNA VAFs varied with progression status. We found no difference between pre- and on-osimertinib CSF-tDNA VAFs among patients with CNS PD (*P* = 0.67) but a trend towards lower CSF-tDNA VAFs among patients without CNS PD (*P* = 0.08, **Fig. 6B**). CSF samples from the trial were also analyzed for osimertinib penetration rate (CSF osimertinib concentration normalized by plasma osimertinib concentration) and we therefore explored the association between drug penetration and on-treatment CSF-tDNA concentration. There was no significant correlation between osimertinib penetration rate and on-osimertinib CSF-tDNA concentration (Spearman’s ρ, 0.32; *P* = 0.30; **Fig. 6C**). Similarly, there was no significant correlation between osimertinib CSF penetration and the difference in on-osimertinib and pre-osimertinib VAF (Spearman’s ρ, 0.32; *P* = 0.46), as was the association between plasma osimertinib concentration and post-osimertinib plasma ctDNA VAF (**Fig. S3A-B**). Finally, we investigated the utility of median CSF-tDNA VAFs, pre-osimertinib and post-osimertinib, for predicting response to therapy. While there was no significant association between pre-osimertinib CSF-tDNA VAFs and CNS progression (**Fig. S4**), on-osimertinib CSF-tDNA VAF was strongly associated with CNS progression on therapy (HR = 6.2, *P* = 0.009, **Fig. 6D**).

**Figure 6.**
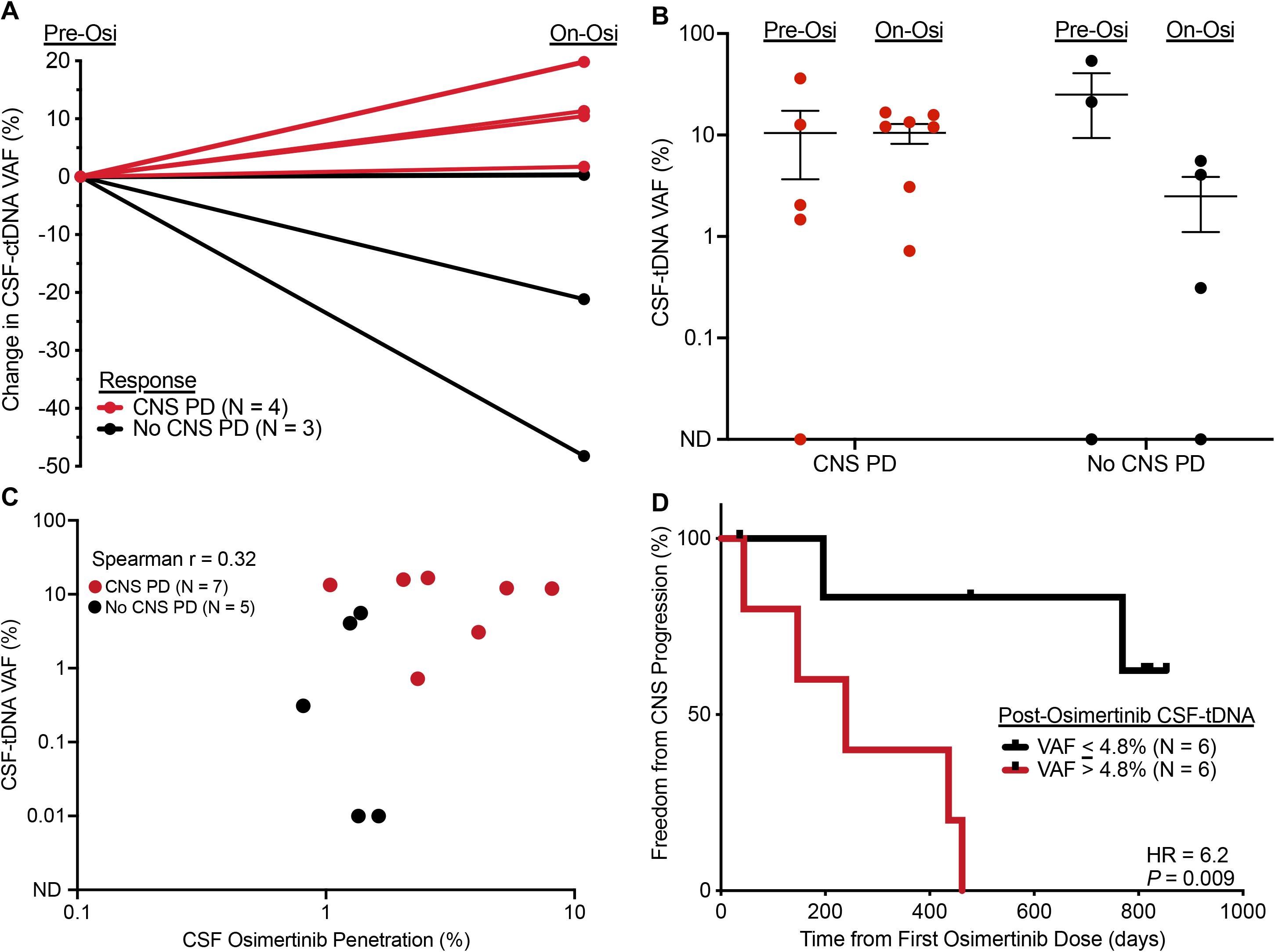
CSF-tDNA detection is associated with progression in the prospective osimertinib cohort. (**A**). Patients with both pre- and on-osimertinib CSF samples available (N = 7), percent change in CSF-tDNA VAF. (**B**) Comparison of CSF-tDNA VAF pre- and post-osimertinib in patients with CNS PD (*P* = 0.67) and patients without CNS PD (*P* = 0.08). P values calculated by the Mann-Whitney test. (**C**) Correlation of CSF-tDNA VAF with percent CSF osimertinib penetration, colored by CNS progression status (red, CNS PD; black, no CNS PD). Percent CSF osimertinib penetration was calculated by normalizing CSF osimertinib concentration (nM) to plasma osimertinib concentration (nM). P value and ρ were calculated by Spearman correlation. (**D**) Kaplan-Meier curve comparing patients with post-osimertinib CSF-tDNA VAF above and below median (4.8%) for endpoint of freedom from CNS progression (*P* =0.009, HR = 6.2 (95%CI, 1.2-31.8)). P value and hazard ratio were calculated from the log-rank test.

## Discussion

In this study we provide evidence for the utility of CSF-tDNA analysis in the management of NSCLC patients with LMD. We found that resistance mutations detected in plasma ctDNA are often absent from CSF-tDNA, suggesting evolution of distinct resistance mechanisms based on the location of tumor deposits. Additionally, we found that CSF-tDNA detection at time of LMD diagnosis and drop of CSF-tDNA concentration in response to targeted therapy with osimertinib appears to have prognostic value, potentially facilitating improved patient counseling and risk stratification.

We observed that detection of CSF-tDNA may be more sensitive than current methods of LMD diagnosis. While CSF cytology is highly specific, it has sensitivity of only 50-60% on initial LP,^1, 9^ and is therefore an imperfect gold standard. In our cohort, CSF-tDNA was 91.7% sensitive. Furthermore, 4/9 samples which were cytology negative had detectable CSF-tDNA in suspected LMD patients.

A well-established barrier to long-lasting effectiveness of molecular therapies is the inevitable emergence of sub-clonal tumor populations harboring resistance mutations. Prior studies in NSCLC have demonstrated spatiotemporal heterogeneity of canonical resistance mutations, potentially offering opportunities for therapeutic re-challenge schemes and selective targeting of specific body compartments.^45^ EGFR-TKI re-challenge remains promising, with multiple studies evaluating re-administration of EGFR-targeted therapy after salvage cytotoxic chemotherapy.^46, 47^ In the selection of patients most likely to benefit from TKI re-challenge, plasma ctDNA and CSF-tDNA could allow personalized surveillance of resistance mechanisms within and outside of the CNS.^27, 31–33, 48^ Early changes in clonal composition, particularly those conferring therapeutic resistance, could aid clinical decision-making in identifying drugs most likely to be effective. Additionally, beyond the dynamic heterogeneity in tumor genotype over time, spatial heterogeneity may also help direct therapy.^48^

Secondary resistance EGFR mutations, such as T790M with 1^st^ and 2^nd^ generation EGFR-TKIs and C797S mutation with 3^rd^ generation EGFR-TKI, are common causes of acquired resistance to EGFR-TKIs in EGFR-mutant lung cancer. However, these secondary EGFR mutations appear less frequently in CNS resistance samples from EGFR-mutant lung cancer patients.^27, 32^ One hypothesis regarding this observation is that limited penetration of targeted therapeutics into the CNS due to the blood brain barrier results in different clonal selection pressures between the CNS and periphery. Another hypothesis is that resistance arises because of the different microenvironments in the periphery versus the CNS, such as due to differences in secreted factors. Consistent with this hypothesis, recent evidence suggests that the EGFR ligand amphiregulin induces resistance in an EML4-ALK rearranged lung cancer LMD mouse model, a finding which was also confirmed in human CSF samples.^49^ Analogous mechanisms could potentially lead to EGFR TKI resistance.

In the subset of our patients who received osimertinib, a third-generation EGFR-TKI demonstrated to have improved CNS response,^50–52^ previously described resistance mutations in LMD were observed, with *MET* copy number gain seen in the CSF-tDNA of 1/5 patients with CNS progression.^53^ We observed that resistance mutations were more prevalent in time-matched peripheral blood than in CSF, consistent with prior reports.^27, 32^ While the mechanisms driving this observation were not assessed in our study, future investigations into this divergence are warranted. A compartment-specific framework may meaningfully inform the development of patient-specific therapeutic decision-making.

### Longitudinal Interrogation of Molecular Response

The assessment of clinical response in patients with LMD remains challenging due to limitations of MRI imaging, the lack of standard evaluation criteria, and the relatively poor sensitivity of cytology.^28, 54^ Notably, none of these methods are quantitative. Several prior studies have demonstrated the potential promise of CSF-tDNA in advanced NSCLC patients with *EGFR* and *EML4-ALK* mutant tumors.^24, 26–28, 31^ Notably, these studies did not include patients from prospective cohorts. In our prospective osimertinib cohort, we found that, while pre-treatment CSF-tDNA levels were not prognostic, mid-treatment CSF-tDNA after only 3 weeks of treatment were elevated in patients who went on to develop CNS progression. This is a key finding for future studies, because it offers an opportunity to adapt therapy early on in patients destined to develop rapid clinical deterioration. For example, one could envision early therapeutic change or escalation (*e.g*., addition of radiotherapy) in patients who do not show a drop of CSF-tDNA concentration at the 3-week mark. Future prospective clinical trials will be required to test the utility of such an approach.

Prior manuscripts in other cancer settings have described the importance of ensemble mutational consideration when quantifying clinical response.^39, 55–57^ Prior explorations of molecular response to osimertinib relied in individual reporters such as CDK4 and EGFR alterations and even the lowest risk groups had a median time to progression of ∼10-15 months. Furthermore, prior studies have near-uniformly relied on the digital readout of whether these individual alterations are detectable.^58^ Such approaches may be susceptible to assay detection limit parameters compared to ensemble approaches for disease quantification. Our prospective cohort receiving osimertinib demonstrated robust risk stratification with the low-risk group not reaching 50% intracranial progression despite over two years of follow-up. Particularly for clinical applications involving therapeutic selection and timing, maximizing risk stratification is crucial particularly for rapidly progressive pathologies like LMD.

Despite our findings, additional work is necessary to more thoroughly understand how spatiotemporal heterogeneity emerges in the context of LMD. Studies of tumor evolutionary history allows for the construction of patient-personalized phylogenetic trees visualizing the accumulation of genomic aberrations over time.^59^ Future studies could recapitulate these findings in parallel analyses of the CNS and systemic compartments in patients with CNS tumor involvement to temporally resolve the differences observed in our study. Additionally, while we describe the landscape of peripheral blood- and CSF-associated genetic aberrations in LMD, additional functional studies are necessary to identify their significance in disease pathogenesis and to elucidate putative druggable targets.

There are some limitations to this study. The first is that the number of CSF samples at the time of resistance was very small because it is not standard practice to do LP at the time of resistance. The second is that the amount of CSF was relatively limited due to potential adverse effects of large volume CSF collection.

In conclusion, we provide evidence supporting the utility of CSF-tDNA detection to diagnose LMD in lung cancer patients. Assessment of CSF-tDNA has the potential to measure molecular response of LMD to EGFR-TKI treatment at early treatment timepoints with implications for therapeutic decision making. The use of dynamic measurement of plasma ctDNA and CSF-tDNA status and profiling of co-occurring gene alterations provides support for prospectively testing novel strategies for personalized management of this devastating clinical entity.

## Materials and methods

### Study cohorts

The samples analyzed in this article were collected at two institutions between 2015 and 2020. Thirteen patients were recruited at Stanford Hospital and Clinics and underwent lumbar puncture as part of routine clinical management of suspected LMD. Samples from twelve patients who were enrolled in a prospective study to determine the efficacy of osimertinib in the treatment of refractory LMD at the Institute of Biomedical Research and Innovation Hospital were also included.^44^ In the prospective cohort, collected and analyzed samples included: a pre-osimertinib sample (CSF, N = 9; tumor, N = 1; pleural effusion, N = 2), a post-osimertinib CSF sample (N = 12), and a time-matched post-osimertinib plasma sample (N = 12). The post-osimertinib samples were collected either at time of disease progression or at last available follow up. This prospective cohort is referred to as the “prospective osimertinib cohort” throughout the manuscript. In both cohorts, MRI was obtained prior to lumbar puncture. Informed consent was obtained from all patients and enrollment of each cohort was approved by the institutional review board at each respective institution. All patient identifiers are anonymized prior to analysis and are not linkable to health data from the patients they represent. Detailed information regarding patient and samples is depicted in **Fig. S1**.

### Sample collection and processing

All plasma, tumor, and pleural effusion samples were analyzed by CAPP-Seq as previously reported.^41, 42^ Peripheral blood was collected in K_2_EDTA tubes and CSF was collected in standard, plastic LP collection tubes. Cell-free DNA was extracted from CSF samples by QIAamp Circulating Nucleic Acid Kit (QIAGEN, Hilden, Germany) using a protocol adapted from the methods defined by Pentsova *et al.*^15^ Briefly, CSF was placed on ice after collection and centrifuged at 1800 x *g* for ten minutes. The supernatant was transferred to a second tube and then centrifuged again at 20,000 x *g* for an additional ten minutes. Cell-free DNA was isolated and used in downstream applications in keeping with our previously described CAPP-Seq methodology,^41, 42^ described briefly below.

### Library preparation and targeted next-generation sequencing

DNA isolation, library preparation and targeted sequencing were performed using iDES-enhanced CAPP-Seq previously described.^41, 42^ Briefly, plasma, CSF, and germline DNA (from peripheral blood mononuclear cells (PBMCs) in plasma-depleted whole blood) were used to build sequencing libraries and subjected to targeted exome capture using a previously published lung cancer CAPP-Seq selector.^60^ Sequencing was performed on Illumina HiSeq 4000 instruments (Illumina, San Diego, CA) using 2 x 150 paired-end reads with custom adapters for sample multiplexing and molecular barcoding. Sequencing reads were mapped to the human genome (build hg19) followed by removal of PCR duplicates and technical artifacts as previously described.^41, 42^ CSF samples were sequenced to a median deduplicated depth of 150X, plasma samples were sequenced to a median deduplicated depth of 1879X, and germline samples were sequenced to a median deduplicated depth of 1099X. Sequencing data were processed using a custom bioinformatics pipeline and SNV, indel and structural variant calling was performed as previously described.^41, 42^ In keeping with our previous work using iDES-enhanced CAPP-Seq,^41^ cell-free DNA sequencing reads were de-duplicated using molecular barcodes, background-polished to reduce stereotyped base substitution errors, and filtered to limit to the selector space.

### Measurement of osimertinib levels

All Cerebrospinal fluid (CSF) samples were collected after 6 ± 2 h from osimertinib administration and plasma samples were simultaneously collected. The CSF and plasma concentrations of osimertinib were measured using liquid chromatography– tandem mass spectrometry (LC–MS/MS). The CSF penetration rate of osimertinib was estimated based on CSF/plasma concentrations.

### Criteria for CSF-tDNA detection

Samples were analyzed for presence of mutations using CAPP-Seq on plasma, CSF, or plasma-depleted whole blood without *a priori* knowledge of tumor mutations, as previously described.^39, 41, 42^ SNPs were excluded via identification in germline or plasma and protein-coding mutations were retained. A joint set of mutations for each patient was then assessed as a group in the each sample, and a Monte Carlo–based tumor DNA detection index was measured to determine significance (index cutoff point of ≤ 0.05), as previously established.^39, 41, 42^ If the detection index was >0.05, plasma ctDNA or CSF-tDNA was classified as not detected at that time point, whereas if it was ≤0.05 it was classified as detected. The sample plasma ctDNA mutant allele fraction was calculated by averaging the mutant allele fractions for all mutations for that patient. All variant calls are listed in **Table S1**.

### Somatic copy number alteration detection

Somatic copy number alterations (SCNAs) were called using a previously described method.^40^ In brief, SCNAs were detected using a z-score based approach which involves a set of background samples to capture region-specific variabilities in depth across the targeted regions. For each gene, we called focal amplifications and deletions using the targeted regions as determined by the lung cancer CAPP-Seq panel.

### Definition of definitive (1) and possible (2A and 2B) leptomeningeal disease

LMD diagnosis was considered definitive under the following conditions:

1. CSF cytology or positive clinical EGFR CSF PCR in the initial LP *OR*
2A. MRI of the brain or spine performed prior to the diagnostic LP with unequivocal evidence of LMD AND
2B. Progressive neurological symptoms consistent with LMD, following exclusion of other possible causes

MRIs were reviewed for evidence of LMD by a board-certified neuroradiologist (M.I). This definition was adapted based on a prior study evaluating a flow cytometry-based method for LMD diagnosis.^61^

### Statistics

Our primary aim was to test the hypothesis that detection of CSF-tDNA is associated with survival, thus our primary outcome was overall survival (OS; event defined as death from any cause). In our study, all mortality events were attributable to LMD. In the prospective osimertinib cohort, we considered an additional survival endpoint, freedom from CNS progression. This was defined by radiographic and neurological progression. Neurological changes were evaluated by the following factors: disorientation (date and time, location, and name), headache, diplopia, blindness, paresthesia, gait disturbance, and grip strength. We also performed the finger-nose test, eye movement test, meningeal sign test, Barre test, and sense of touch test. Extra-CNS response was evaluated according to the Response Evaluation Criteria in Solid Tumors (RECIST) version 1.1. As CNS radiologic changes are difficult to assess by the RECIST, they were evaluated as improved, stable, and progressed based on findings of dura mater thickening, exuding contrast agent, ventricular distention, and/or, concomitant substantial brain metastases with confirmation by at least two doctors. Time-to-event analysis for survival endpoints was done using the log-rank test to estimate both P values and hazard ratios and expressed as Kaplan–Meier plots. Comparisons of two groups were tested using the Mann-Whitney test. Strength of correlations between continuous variables was assessed using Spearman’s correlation coefficient. All statistical analyses were done using Prism 7 (GraphPad Software) or R v3.2.2 (http://www.r-project.org) through the RStudio environment.

## Funding and Acknowledgements

This work was supported by grants from the National Cancer Institute (M.D. and A.A.A., R01CA188298 and R01CA254179), the US National Institutes of Health Director’s New Innovator Award Program (M.D.; 1-DP2-CA186569), the Virginia and D.K. Ludwig Fund for Cancer Research (M.D. and A.A.A.), the CRK Faculty Scholar Fund (M.D.), the Doris Duke Charitable Foundation (T.D.A.), the Japan Society for the Promotion of Science KAKENHI (S.N. 21K15546 and 23K06648) and Japanese respiratory society fellowship (S.N.). Visualizations created with BioRender.com.

## Data and Code Availability

Reasonable requests for additional code and data access will be reviewed by the senior authors to determine whether they can be fulfilled in accordance with privacy restrictions. Requests for additional materials related to this work should be directed to M.D. and T.D.A.

## Author contributions

Study concept and design: TDA, SN, MD

Acquisition of data: TDA, SN

Analysis and interpretation of data: TDA, SN, MCJ, MD

Drafting of the manuscript: All authors

Critical revision of the manuscript: All authors

Technical and material support: SN, AAA, MD, MHG

Study supervision: AAA, MD

## Data Availability

All data produced in the present work are contained in the manuscript

