## Supplementary figures and images for "Quantification of Cerebrospinal Fluid Tumor DNA in Lung Cancer Patients with Suspected Leptomeningeal Carcinomatosis"

### Fig. S1

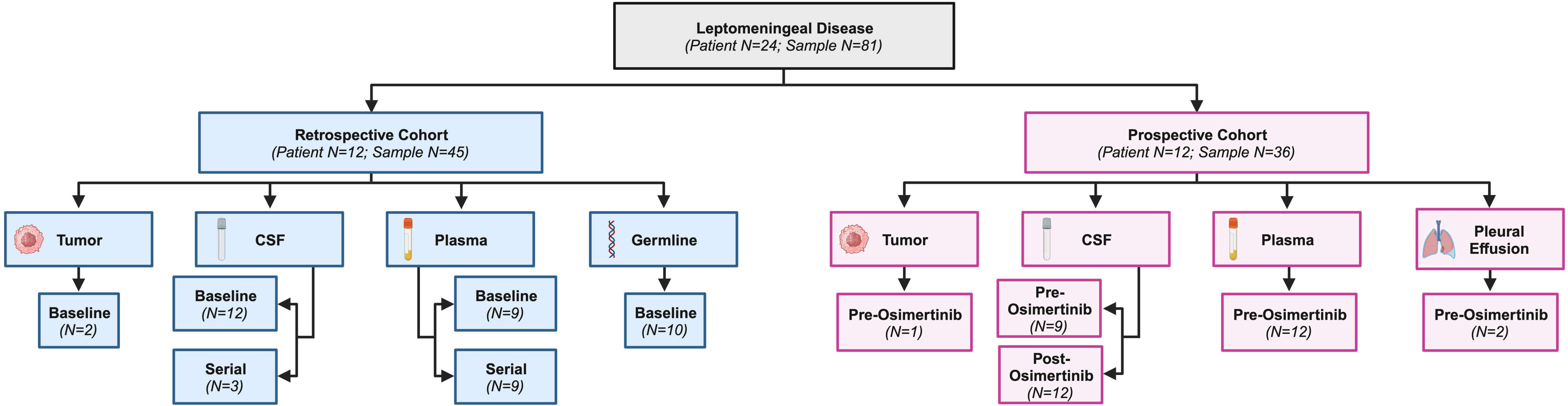

### Fig. S2

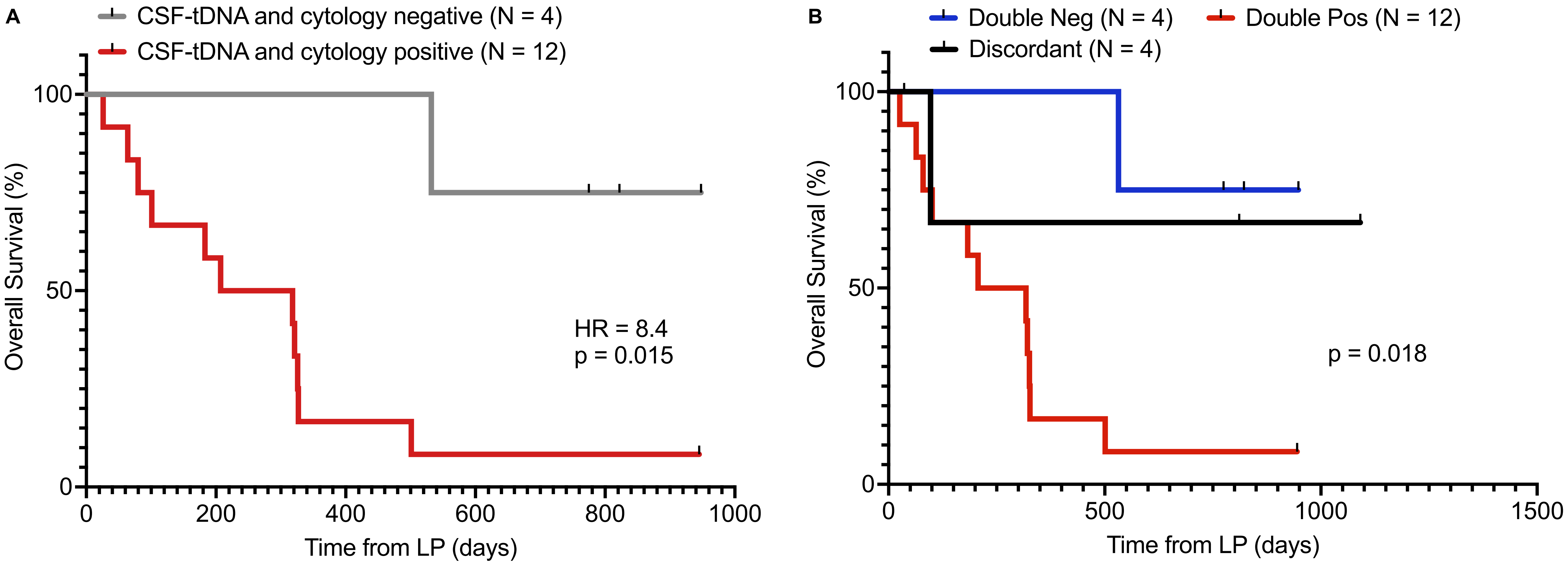

### Fig. S3

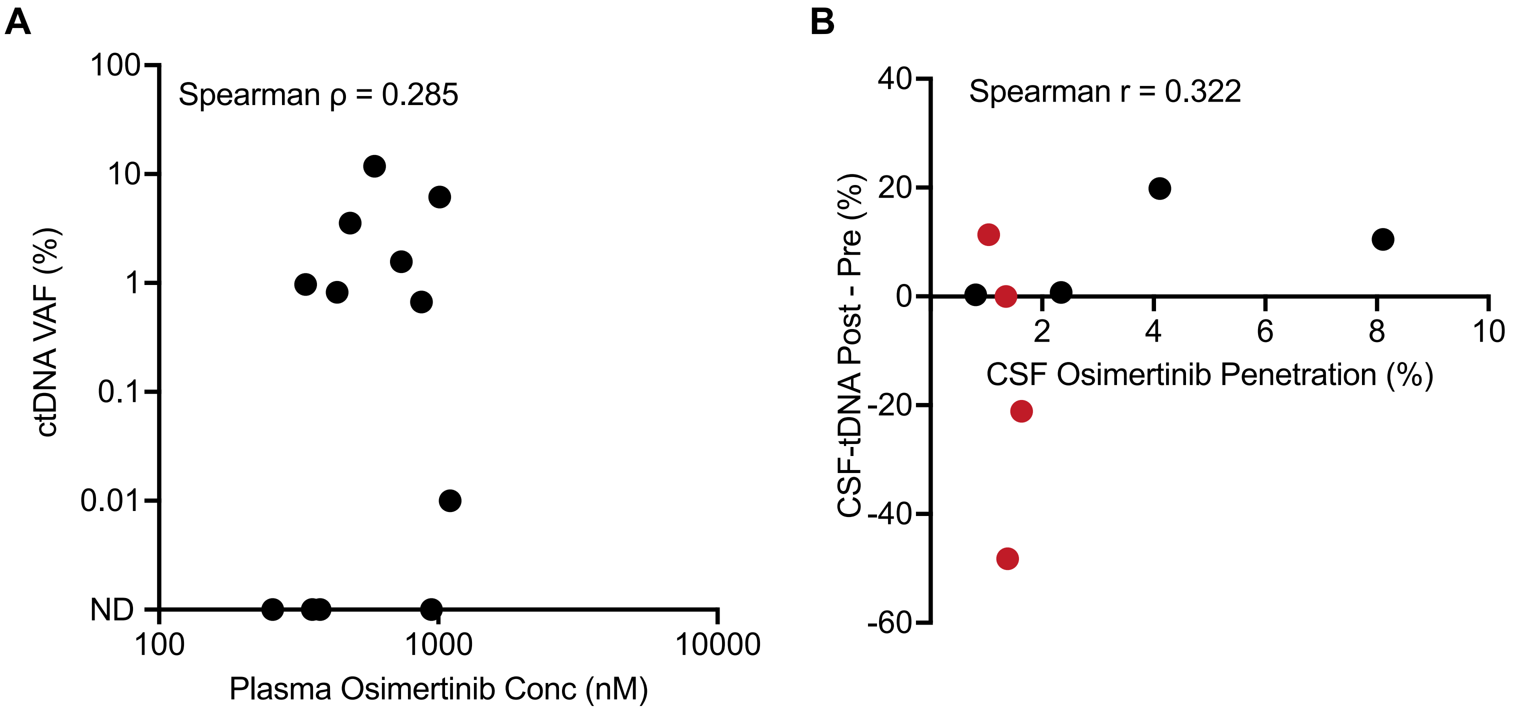

### Fig. S4

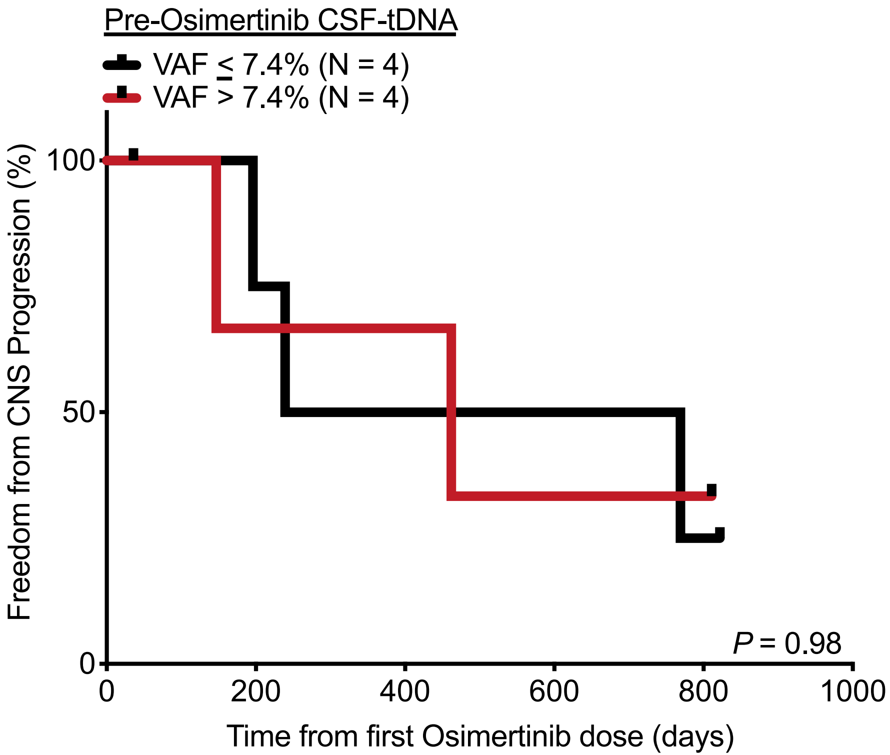
