## Supplementary Legends for "Quantification of Cerebrospinal Fluid Tumor DNA in Lung Cancer Patients with Suspected Leptomeningeal Carcinomatosis"

**Figure S1**. Schematic overview of study patients and samples.

**Figure S2**. (**A**) Kaplan-Meier curve comparing patients with detectable CSF-tDNA and cytology positive (N = 12) and undetectable CSF-tDNA and cytology negative (N = 4) at first lumbar puncture for the endpoint of overall survival (*P* = 0.008, HR = 8.6). P value and hazard ratio were calculated from the log-rank test. (**B**) Kaplan-Meier curve comparing patients with CSF-tDNA and cytology negative (Double neg, N=4), patients with CSF-tDNA and cytology positive (Double pos, N=12), and patients with CSF-tDNA positive and cytology negative (Discordant, N=4) for the endpoint of overall survival (*P =* 0.015). P value calculated from the log-rank test.

**Figure S3**. (**A**) Correlation of post-osimertinib ctDNA VAF %) with plasma osimertinib concentration (nM). ρ was calculated by Spearman correlation. (**B**) Correlation between osimertinib CSF penetration and the difference in on-osimertinib and pre-osimertinib VAF. ρ was calculated by Spearman correlation

**Figure S4.** Kaplan-Meier curve comparing patients with pre-osimertinib CSF-tDNA VAF above and below median (7.4%) for endpoint of freedom from CNS progression (*P* =0.98). P value calculated from the log-rank test.
